# Compliance and containment in social distancing: mathematical modeling of COVID-19 across townships

**DOI:** 10.1101/2020.06.01.20119073

**Authors:** Xiang Chen, Aiyin Zhang, Hui Wang, Adam Gallaher, Xiaolin Zhu

**Author notes:** Corresponding author ORCiD: 0000-0001-6967-786X.

## Abstract

In the early development of COVID-19, large-scale preventive measures, such as border control and air travel restrictions, were implemented to slow international and domestic transmissions. When these measures were in full effect, new cases of infection would be primarily induced by community spread, such as the human interaction within and between neighboring cities and towns, which is generally known as the meso-scale. Existing studies of COVID-19 using mathematical models are unable to accommodate the need for meso-scale modeling, because of the unavailability of COVID-19 data at this scale and the different timings of local intervention policies. In this respect, we propose a meso-scale mathematical model of COVID-19 using town-level infection data in the state of Connecticut. We consider the spatial interaction in terms of the inter-town travel in the model. Based on the developed model, we evaluated how different strengths of social distancing policy enforcement may impact future epidemic curves based on two evaluative metrics: compliance and containment. The developed model and the simulation results will establish the foundation for community-level assessment and better preparation for COVID-19.

---

The year 2020 was deemed to be unprecedented in human history because of the outbreak of the novel and infectious coronavirus (COVID-19). As of early May 2020, the virus has led to over 3.5 million confirmed infections, over 250 thousand deaths, and echoes of economic depression around the globe (Dong, Du, and Gardner 2020). The United States has become the largest victim in this public health calamity. The effect of the virus latency, coupled with the lack of clinical interventions, was further amplified by the understatement of the disease severity in its early development in the country. After COVID-19 was declared by the presidential proclamation as a national emergency on March 1, 2020 (White House 2020), many state and local governments started to enforce strict preventive measures to mitigate the community spread (Parmet and Sinha 2020).

These preventive measures, known as social distancing (e.g., closure of non-essential businesses, stay-at-home order), aim to minimize interpersonal interactions (Gostin and Wiley 2020). It has been found that these measures have been effective in delaying the spread of the virus by flattening the epidemic curve (epi curve) through the observation of transmission (Anderson et al. 2020). While early discussion of social distancing revolved around social impacts such as the economic consequences (Atkeson 2020) and ethical paradoxes (Lewnard and Lo 2020), many recent studies have integrated social distancing into mathematical epidemic models, attempting to simulate and predict scenario-based future outbreaks (Chen et al. 2020; Kissler et al. 2020). These models, however, have been largely focused on the macro-scale using a relatively large geographic unit, such as country (Gilbert et al. 2020; Kissler et al. 2020), state (Chen et al. 2020), or county (Lai et al. 2020). To the authors’ knowledge, there have been no epidemic models investigating the COVID-19 development at the meso-scale with a smaller geographic unit, such as town or census tract.

## 1. The scale issue in modeling COVID-19

The meso-scale, generally known as a study area of 1–1000 kilometers, is of critical importance in the effective containment of the epidemic growth. This significance can be justified by the mechanism of the preventive strategies over the different phases of epidemic development. In the early development of COVID-19, large-scale preventive measures, such as border control and air travel restrictions, were implemented to slow international and domestic transmissions. When these measures were in effect, new cases of infection would be primarily induced by community spread, such as the human interaction within and between neighboring cities, towns, and communities. Existing macro-scale studies using classical epidemic models, notably the Susceptible, Exposed, Infectious, Recovered (SEIR) model are unable to accommodate the need for meso-scale modeling, because of three existing limitations in COVID-19 research.

First, in the United States, the timing of the COVID-19 outbreak differs by state and so do their regulatory countermeasures, such as the enforcement of the stay-at-home order. It is relatively intractable to model COVID-19 at a macro-scale while considering the heterogeneity in the timing of local policies and the strength of their enforcement. Second, when long-distance travel (e.g., flights) are restricted, the transmission will be dictated by short-distance travel, such as daily commuting trips by public transit or private automobiles. In this context, individual mobility and the likelihood of travel is largely driven by the compliance of social distancing rules. Therefore, modeling COVID-19 at the meso-scale should articulate how social distancing affects people’s travel activities or the willingness to travel as parameters to model the process of transmission. This gap has not been fulfilled by the status quo macro-scale models. Third, while COVID-19 data (e.g., infection, death, and recovery) on a daily basis has become largely available in the public sector, data with finer spatial granularities, such as across townships or census tracts, are extremely lacking. These three tiers of research gaps fuel the need to develop a meso-scale epidemic model that simulates past COVID-19 cases while predicting the local, community-level spread in preparing for a highly likely resurgence in the near future.

In this paper, we propose a meso-scale epidemic model using town-level COVID-19 infection data in the state of Connecticut. Because the local infection is largely subject to effects of social distancing, the model development follows two evaluative metrics in social distancing: compliance, which represents the strength of the policy enforcement, and containment, which represents individual mobility. By incorporating these two metrics into the SEIR model, we have proposed a meso-scale SEIR model and have performed model fitting and sensitivity analysis under ten different social distancing scenarios. Using the developed model, we have evaluated how different social distancing strategies (i.e., minimal, moderate, and substantial) would shape the epi curve of COVID-19 for each town. This study, while among the first to simulate the COVID-19 development at the meso-scale, has the potential to inform both epidemiologists and stakeholders about the public health risks using different social distancing strategies.

The paper is organized as follows. Following the background, Section 2 introduces the methodological development of the model based on the classical SEIR model and the guiding principle of social distancing. Section 3 applies the new model to a case study in Connecticut, performs the model fitting, and simulates epi curves and spatial patterns at the town level based on different social distancing scenarios. Section 4 discusses the major findings and insights shed by the modeling results. Lastly, Section 5 concludes the study with long-term impacts.

## 2. A Meso-scale SEIR model (MSEIR)

### 2.1 Introduction to SEIR model

Our proposed model stems from the classical SEIR model, a deterministic mathematical model to simulate epidemiologic dynamics, as shown in Equations (1) through (4). The SEIR model is composed of four variables: *S* (susceptible population), *E* (exposed population), *I* (infectious population), and *R* (recovered population). It explicitly quantifies a four-stage cycle of the disease spreading among humans in terms of differential equations. Each stage is formulated as a derivate of the population (i.e., *S, E, I, R*) with respect to time (*t*), representing the change of the stage-specific population.

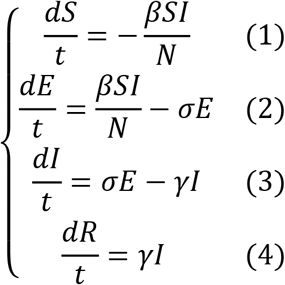

In the equations, *N* denotes the total population (*N* = *S* + *E* + *I* + *R*); *β*, σ, and *γ* are the daily transmission rate, daily incubation rate, and daily recovery rate, respectively. The basic reproduction rate can be derived as R_0_ = *β*/*γ*. This classic SEIR model, along with its many extensions, has been widely applied to epidemic modeling of COVID-19 (Chen et al. 2020, Kissler et al. 2020, Lai et al. 2020).

### 2.2 A Conceptual model of social distancing

Adapting the SEIR model to the meso-scale should emphasize the effectiveness of social distancing in communities. This evaluation follows the Centers for Disease Control and Prevention (CDC)’s social distancing guidelines for COVID-19 given in three aspects: operations of public facilities, restrictions on businesses, and restrictions on personal movement (Gostin and Wiley 2020). For the restrictions on personal movement, the guidelines impose limitations on people’s travel and social behaviors in terms of prohibiting mass gatherings, requiring physical distancing in face-to-face interaction, and enforcing stay-at-home orders (Gostin and Wiley 2020). The CDC also calls for legal and community efforts to enhance compliance with these social distancing measures.

Under this guiding principle, we propose a conceptual model in measuring the effectiveness of social distancing with an emphasis on travel activities as part of the personal movement. The model comprises two metrics, compliance and containment, that evaluate the effectiveness of the policy enforcement, as shown in Figure 1. Compliance evaluates the likelihood of the residents not following the social distancing rules. While there are various ways in which compliance can be articulated, one variable could be the percentage of residents engaging in travel activities as a surrogate for the metric. Containment evaluates the level of human mobility, and a variable for the evaluation could be the maximum distance that people are willing to travel under the social distancing regulation. These two metrics, resulting from the strengths of the policy enforcement, will likely affect the transmission risk of the epidemic. This conceptual model featuring the two evaluative metrics is integrated into the classical SEIR model to develop a meso-scale epidemic model for COVID-19.

**Figure 1.**
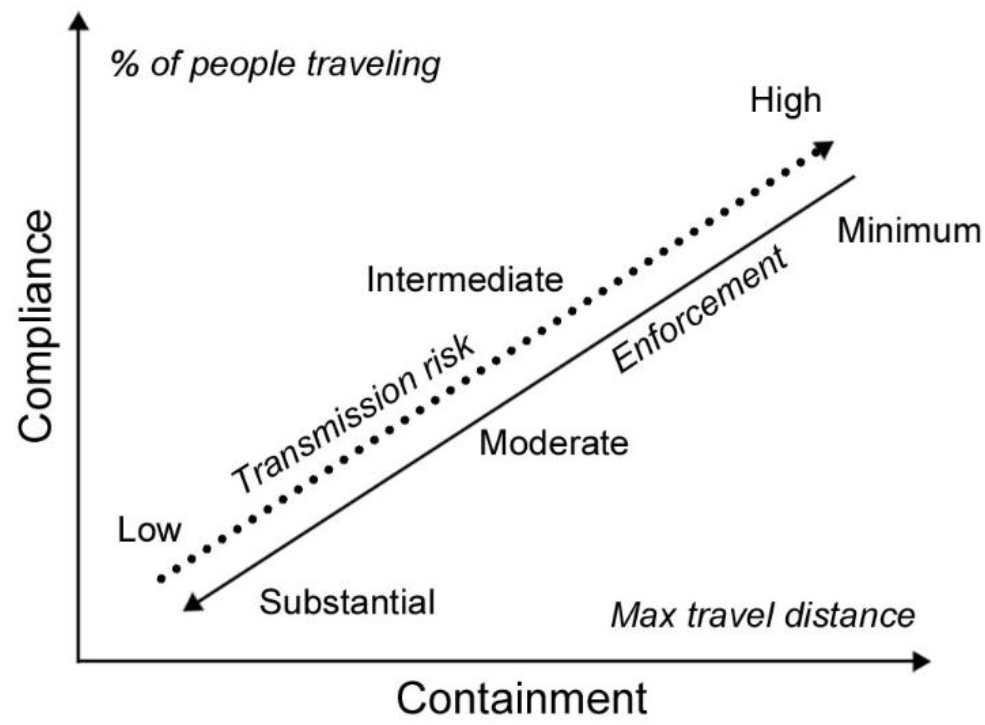
A conceptual model of the effects of social distancing policy on travel activities. The solid line represents the strength of policy enforcement; the dashed line represents the level of transmission risk.

### 2.3 Model development

We modify the SEIR structure with an emphasis on the impacts of travel activities at the meso-scale, where the study area is a state and the unit of analysis is a town (formally known as county subdivision in the United States). Recent epidemic models have employed various forms of mobility data, such as smart-phone heat maps (Lai et al. 2020) and air traffic flow (Gilbert et al. 2020), to estimate mobility in the SEIR model. Because of the lack of mobility data at the town level, we employ the Huff-model (Huff 1963) to estimate the potential for travel. The huff-model is traditionally used for analyzing the business potential based on the probability of customers’ visits to retail and service facilities. It has also been extended to forecasting the external trips between communities (Anderson 1999; Anderson 2005). The classical Huff-model uses real-world survey data to calibrate the model parameters, including the attractiveness of facilities and the distance decay (Huff and McCallum 2008). In this paper, we choose the linear form to estimate the probability of travel between two towns, as shown in Equation (5). This linear form is seen as a better form to forecast inter-city trips and has been corroborated with field data (Anderson 1999).

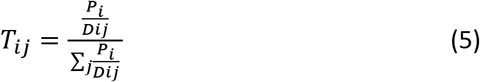

In Equation (5), *T*_ij_ is the probability that a person traveling from town *i* to town *j; D*_ij_ is the distance between *i* and *j*; and *P*_i_ is the population of town *i*.

We have further added to the Huff model two other parameters: a compliance parameter *C*_i_, meaning the percentage of the population of town *i* engaging in inter-town trips, and a containment parameter *D*_0_, meaning the maximum distance people are willing to travel under influences of social distancing. These two parameters extend the Huff-model to estimate *M*_ij_, the total population traveling from town *i* to town *j*, as shown in Equation (6).

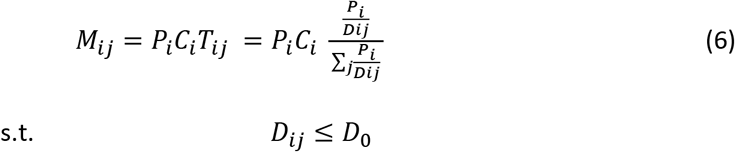

Model (6) is a Huff-based trip distribution model where the compliance parameter *C*_i_ and the containment variable *D*_0_ can be determined by different social distancing scenarios. To further incorporate the trip distribution model to the SEIR model, we have made several necessary assumptions: (1) both the susceptible and exposed populations conform to the mobility rule in Equation(6); (2) the infectious and recovered populations are isolated so that they cannot travel to other towns; (3) the susceptible population traveling to other towns can be affected by the infectious population in both their origin town and destination town; (4) the total population and the daily travel population of a town are stable during the modeling period; (5) daily travelers return to their origin town by the end of the day; and (6) the transmission rate gradually decreases due to non-pharmaceutical interventions during the social distancing period (Lai et al. 2020). Based on these assumptions, we have developed the meso-scale SEIR model (MSEIR), simulating the daily dynamics of susceptible (*S*_i_), exposed (*E*_i_), infectious (*I*_i_), and recovered populations (*R*_i_) of the *i*th town, as shown in Equations (7) through (11).

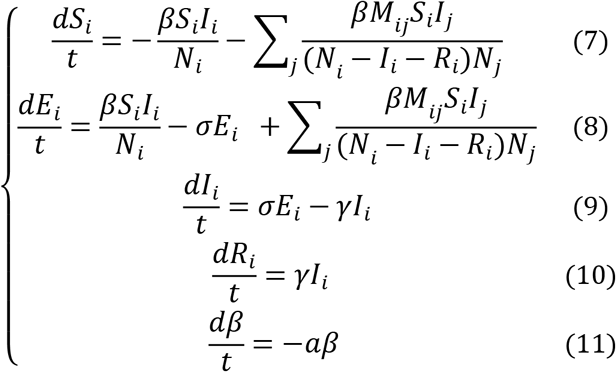

where

*E*_i_: exposed population of town *i*;
*I*_i_: infectious population of town *i*;
*M*_ij_: population (susceptible or exposed) traveling from town *i* to town *j*. The parameter is derived from Equation (6).
*N*_i_: total population of town *i* (*N*_i_ = *S*_i_ + *E*_i_ + *I*_i_ + *R*_i_);
*R*_i_: recovered population of town *i* (including hospitalized, self-recovered, and death);
*S*_i_: susceptible population of town *i*;
*t*: time (daily);
*a*: daily change rate of the transmission rate (0 < *a* < 1);
*β*: transmission rate (conversion from the susceptible population to exposed population);
*γ*: recovery rate;
σ: daily incubation rate (reciprocal of the incubation period);

In the MSEIR model, Equations (7) through (10) are a series of differential equations indicating the daily variation of the susceptible, exposed, infectious, and recovered population at each transmission stage. Equation (11) is the daily change of the transmission rate.

### 2.4 Model initialization and parameter estimation

To implement the model, five state variables at the initial stage must be derived: (1) the initial transmission rate *β*_0_ (estimated from our case study, see below); (2) the initial exposed population *E*_i0_ (estimated from our case study, see below); (3) the initial infectious population *I*_i0_, which equals to the cases of infection on March 23, 2020, the start date of the data. (4) the initial susceptible population *S*_i0_, which can be derived as *S*_i0_ = *N*_i_ – *E*_i0_; and (5) the initial recovered population *R*_0_ = 0 under the assumption that no individuals are cured, hospitalized, or had died at the initial stage.

*σ* and *γ* can be derived from historical data. *σ* is the daily incubation rate as the reciprocal of the incubation period. *γ* is the recovery rate, indicating the rate of reduction in the infectious population due to hospitalization, self-recovery, and death. This parameter assumes that once an infectious individual is hospitalized, self-recovered, or died, the person will be isolated from the transmission cycle. According to a recent study among the first 425 diagnosed patients (Li et al. 2020), the mean incubation period of COVID-19 was 5.2 days (at a 95% confidence interval [CI], 4.1 to 7.0 days), and the mean time duration from the illness onset to hospital admission was 9.1 days (95% CI, 8.6 to 9.7 days). Thus, we assumed that our study had a similar incubation period and duration from illness onset to the first medical visit. In our model, *σ* = 1/5.2 and *γ* = 1/9.1.

For parameters *β*_0_ and *a*, we estimated their optimal values for each town using the daily cumulative cases of infection derived from the Connecticut Department of Public Health (CDPH, 2020). We assumed that *β*_0_ and *a* were constant throughout the study area and under different social distancing scenarios. The exposed population at the initial stage *E*_i0_ was an unknown parameter that varied by town. Because of this uncertainty, we treated *E*_i0_ as another parameter to be estimated. Thus, each town *i* had an independent *E*_i0_ given the difference in the onset of the outbreak, population, and other factors dictating the early exposed population. The Nelder-Mead algorithm (Nelder and Mead 1965) was employed to estimate parameters by minimizing the negative normal log-likelihood between the simulated and the confirmed daily cumulative cases.

## 3. Case study

### 3.1 Study area and data

Our study was implemented in the state of Connecticut with the unit of analysis being county subdivision or town. Located in the New England region, Connecticut is the third smallest state by area in the United States with 169 towns and a total population of 3.5 million (Figure 2a). On March 8, the first COVID-19 case was reported in Wilton, a town neighboring New York (The New York Times, 2020). Because of the geographical proximity to New York City, the epicenter of the national outbreak, the state experienced an exponential rise of infections in the early outbreak. As of May 11, the total confirmed cases of infection were over 34,000, and the total deaths were over 3,000 (CDPH 2020). Figure 2b shows the COVID-19 infection rate per 10,000 people as of May 11, 2020 (Figure 2b).

**Figure 2.**
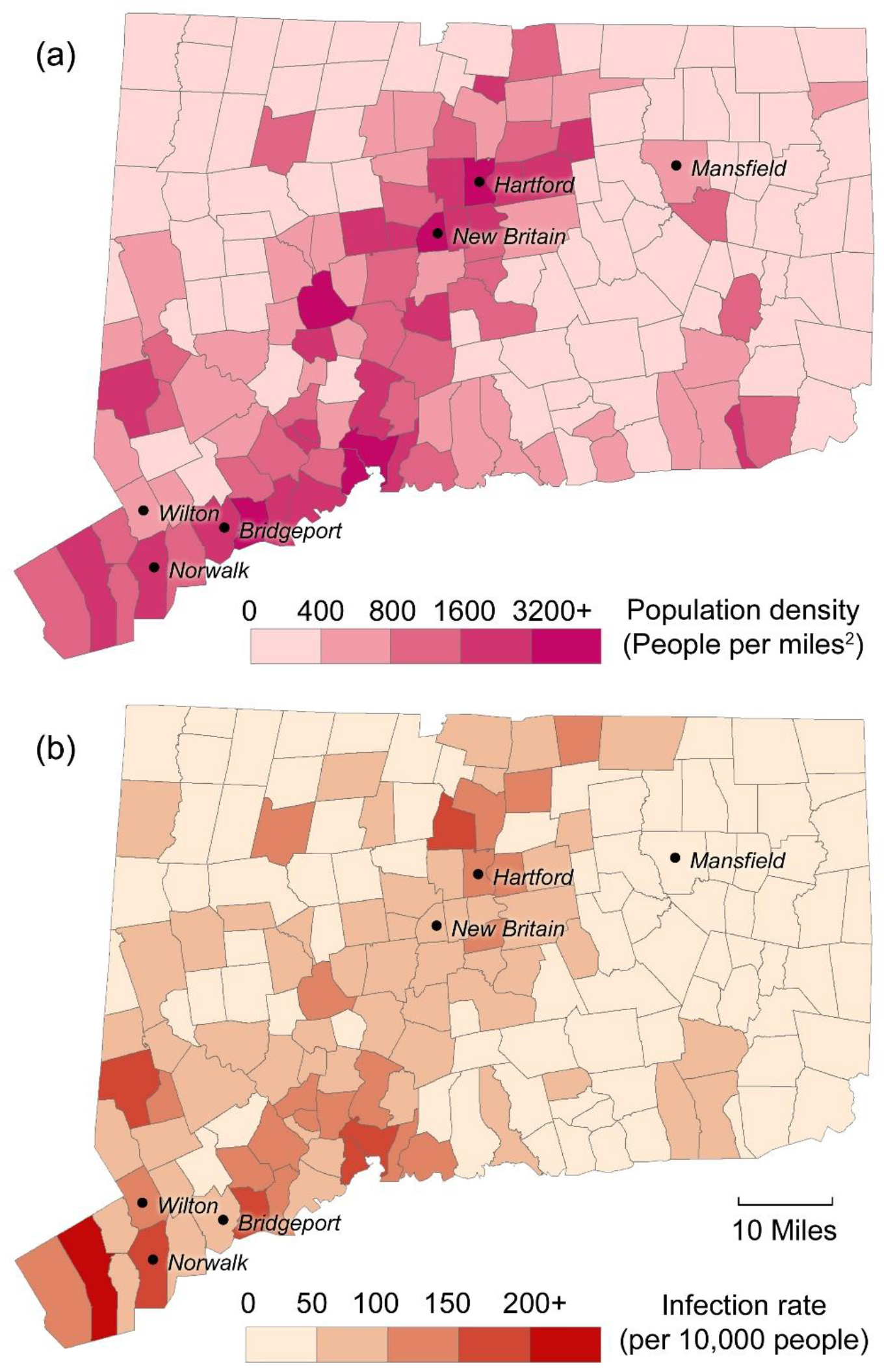
Connecticut towns with (a) population density of 2018 and (b) COVID-19 infection rate as of May 11, 2020. Towns further discussed in the article are labeled.

Modeling COVID-19 at the meso-scale is inseparable from state policy related to social distancing. In Connecticut, the first stage of the social distancing rules was implemented on March 23 by the governor’s executive order “Stay Safe, Stay Home,” requiring the closure of non-essential businesses and some non-profit organizations (Ct.gov 2020a). This policy was relieved by reopening certain non-essential businesses effective on May 20 (Ct.gov 2020b). Based on this context, we retrieved the daily town-level COVID-19 infection data during this period, specifically the first 50 days since the first day of enforcing the state social distancing rules (i.e., March 23 through May 11, 2020). The dataset was solicited from the state government’s daily publications (CDPH 2020).

### 3.2 Social distancing scenarios

We have designed three compliance levels and three containment levels to estimate *M*_ij_ in the MSEIR model, forming a total of nine models representing different degrees of social distancing policy enforcement, as shown in Table 1. In this framework, Model 1 represents the substantial enforcement with only 10% of the population taking inter-town trips and a maximum travel distance of 20 miles; Model 9 represents the minimum enforcement with 50% of the population taking inter-town trips and a maximum travel distance of 140 miles. The 140-mile threshold is the road network distance between the two most remote towns in Connecticut (i.e., Thompson and Greenwich).

**Table 1.** Nine social distancing scenarios based on different compliance and containment levels.

| Compliance level ($C_i$) | Containment level ($D_0$) | | |
| --- | --- | --- | --- |
|  | 20 miles | 60 miles | 140 miles* |
| 10% | Model 1<br>(Substantial) | Model 2 | Model 3 |
| 30% | Model 4 | Model 5<br>(Moderate) | Model 6 |
| 50% | Model 7 | Model 8 | Model 9<br>(Minimum) |

These social distancing scenarios were incorporated into Equation (6) by Python scripting to derive *M*_ij._ In the implementation, *P*_i_ was derived from the 2018 census data; *D*_ij_ was derived as the road network distance between the geographic centers of towns using the Network Analysis module in ESRI ArcMap 10.7 (i.e., OD cost matrix) in a refined road network. The results of M_ij_ were imported to the MSEIR model (scripted in Python) for fitting the dynamics of COVID-19 infection in the 50-day period. We then employed the fitted models for predicting future trends.

### 3.3 Results

Using the town-level data, we implemented the MSERI model under all social distancing scenarios in Table 1. In addition to the model fitting, we extended the epi curves of the cumulative cases of infection (*I_c_*) to the future development with an end date of July 12, 2020. We also established the baseline scenario in the SEIR model where there is no travel or interaction between towns (*C*_i_ = 0%, *D*_0_ = 0 miles).

For the sake of clarity, we selected four scenarios for comparison and discussion: the SEIR model for no interaction, Model 1 for substantial social distancing, Model 3 for moderate social distancing, and Model 9 for minimum social distancing. Figure 3 shows the COVID-19 epic curves of the entire state. As shown in the figure, the SEIR model has considerably underestimated the epi curve, with the value of *I_c_* converging to 26,000. When the three social distancing scenarios are introduced, the epic curves start to align with the confirmed cases, with the minimum social distancing scenario (Model 9) yielding the steepest curve. The simulation predicts that the total cumulative cases of infection will be in the range of 45,752–48,105 (Model 5 and Model 9) as of July 12, which is a rise by 34.2%–41.1% with respect to the reported cases (*I_c_* = 34,070) as of May 11.

**Figure 3.**
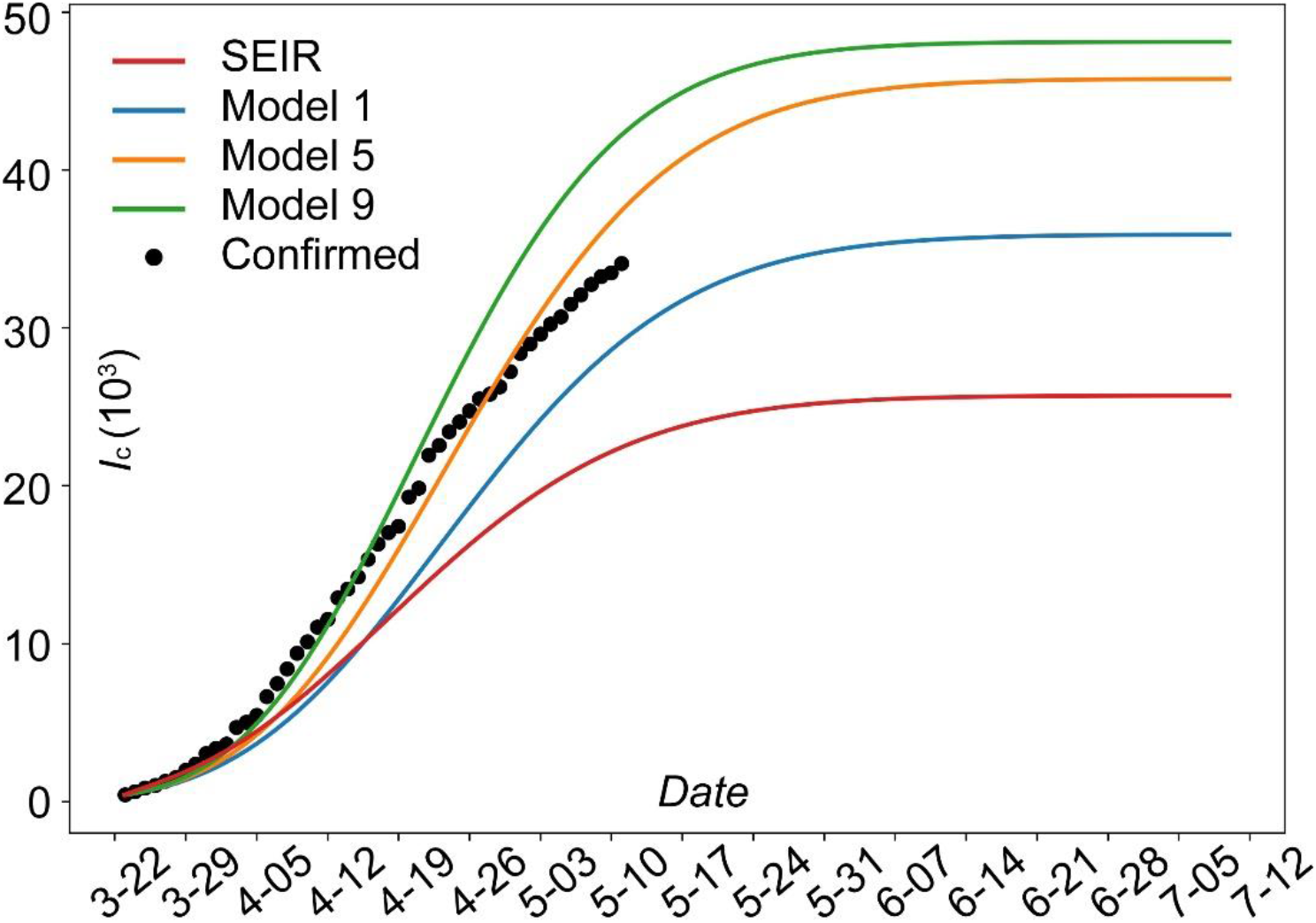
Simulation results of the cumulative cases of infection (*I*_c_) by the SEIR model and the MSEIR model (Models 1, 5, and 9) in Connecticut.

Figure 4 shows the COVID-19 epic curves for six selected towns. As shown in Figure 2, Bridgeport (population: 144,900), Norwalk (population: 89,047), and Wilton (population: 18,397) are located in the center of outbreak in Southwest Connecticut; the capital city Hartford (population: 122,587) and its satellite city New Britain (population: 72,453) are located in another high-risk region; Mansfield (population: 70,981) is a relatively rural area comprised of mainly university employees and students. It can be seen from the results that the SEIR model with no spatial interaction generates the flattest curve for all six towns and underestimates the infection in Bridgeport, Hartford, New Britain, and Norwalk. However, the results under different social distancing scenarios are mixed, and all simulation results for Hartford are somewhat under-estimated. We feel these uncertainties could be a result of the discrepancies between the simulated travel activities and the real-world travel flow.

**Figure 4.**
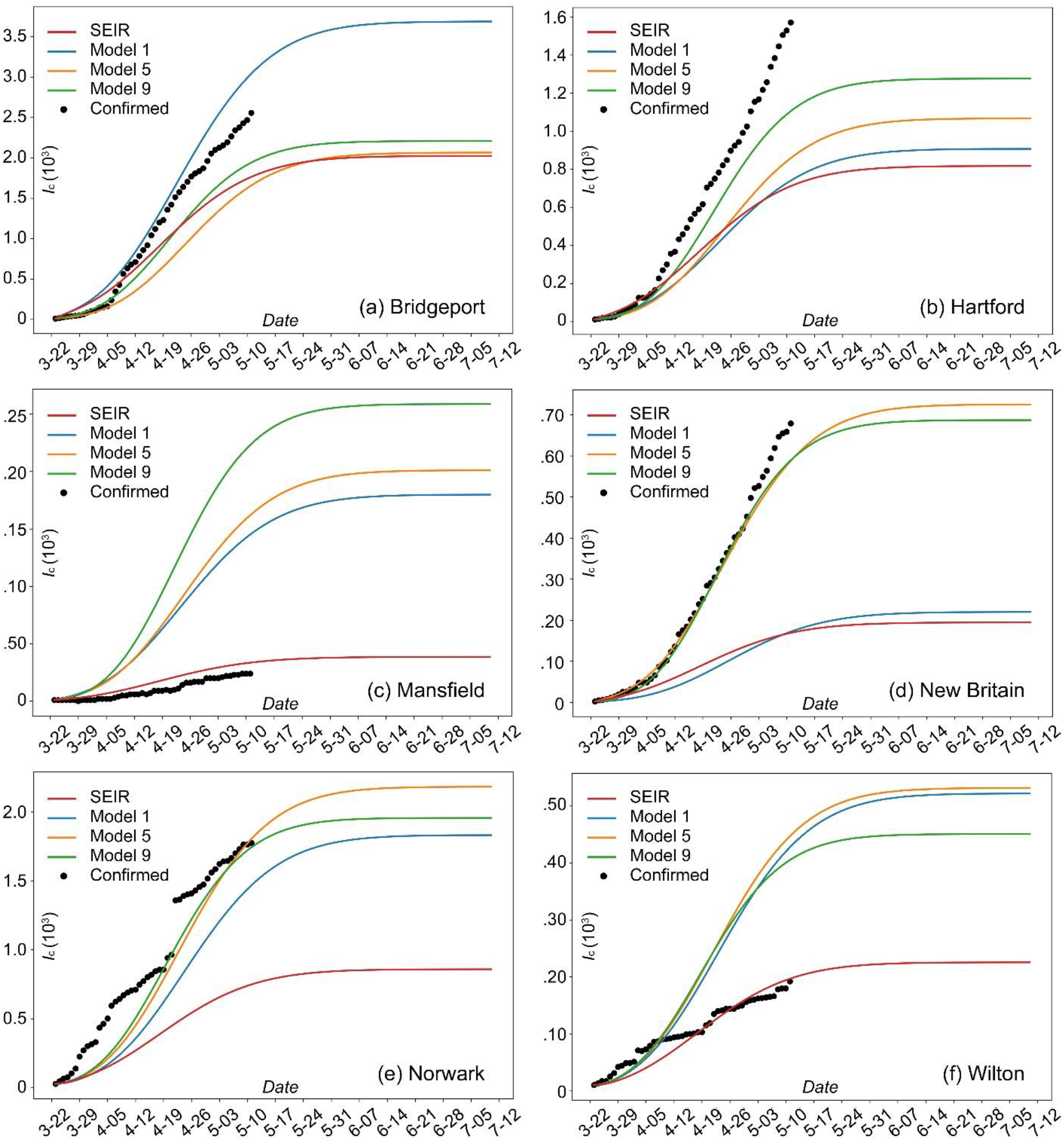
Simulation results of the cumulative cases of infection (*I*_c_) by the SEIR model and the MSEIR model (Models 1, 5, and 9) for six Connecticut towns: (a) Bridgeport, (b) Hartford, (c) Mansfield, (d) New Britain, (e) Norwalk, and (f) Wilton. The geographical locations of these towns are shown inFigure 2.

Figure 5 is a comparison of the simulated infection rates across all towns as of July 12. The map data were derived from the SEIR model, Model 1, Model 5, and Model 9. Results for other models are given in the Appendix. As all the figure illustrates, towns with the highest infection rates are located in Southwest Connecticut, which is a pattern consistent with the existing outbreak. In addition, substantial social distancing measures (Model 1, Figure 5b) can largely curb the local spread, comparing to the moderate (Model 5, Figure 5c) and the minimum control scenarios (Model 9, Figure 5d). We then scrutinized the results by deriving the rate of increase per 10,000 people for each town between May 11 and July 12 under each of the ten models. Then, we counted the frequency of the town appearing on the top 10 list. We found that the following towns appeared on the list at least five times: Westport (*N* = 10), Bethlehem (*N* = 10), Stafford (*N* = 6), New Canaan (*N* = 6), Darien (*N* = 6), Ridgefield (*N* = 6), Wilton (*N* = 5), and Weston (*N* = 5). These towns, as labeled in Figure 5, have a higher likelihood of outbreak during May, June, and July.

**Figure 5.**
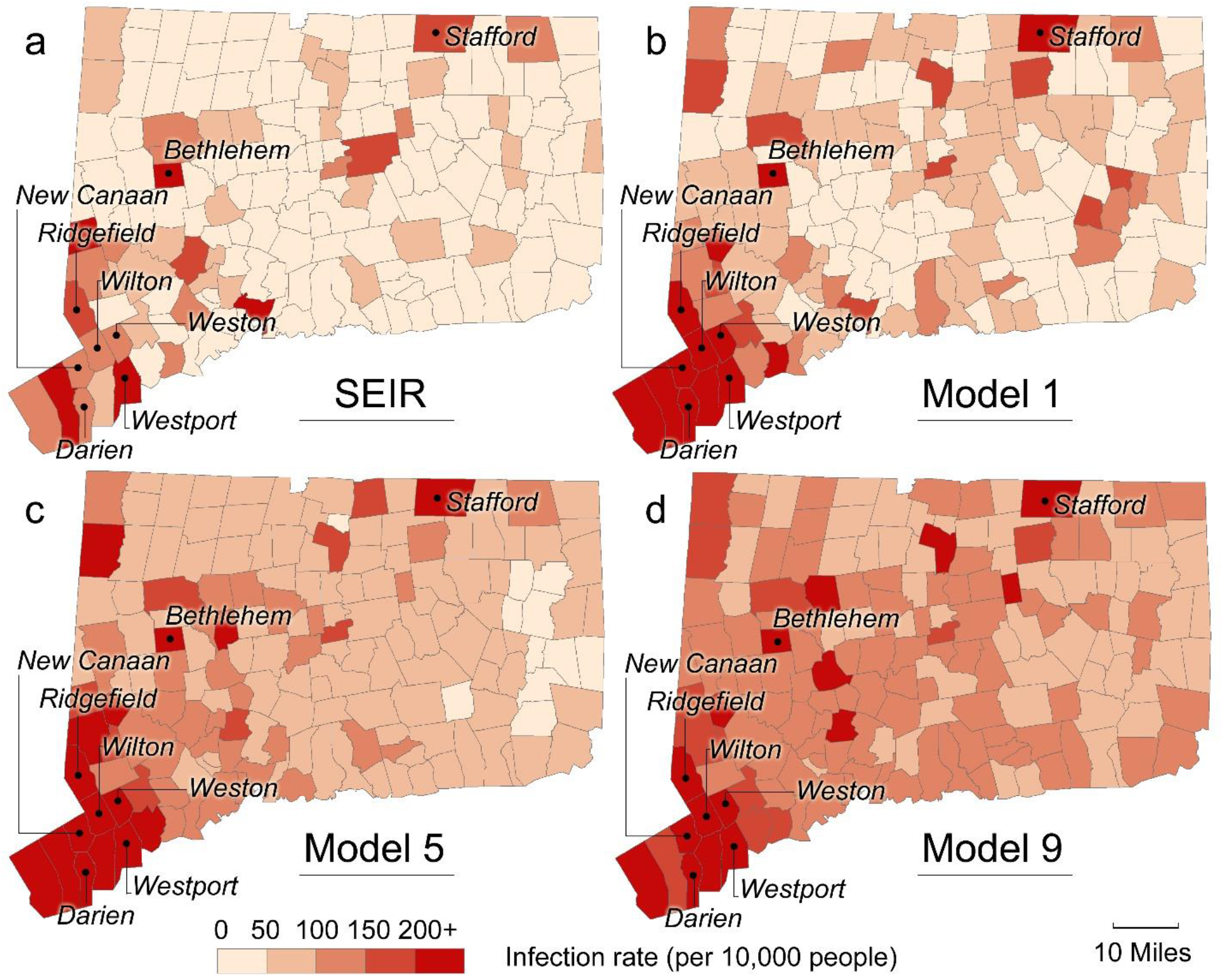
Simulated COVID-19 infection rates (per 10000 people) as of July 12 based on (a) SEIR model, (b) Model 1, (c) Model 5, and (d) Model 9. Towns with the highest rate of increase (appearing on the top 10 list for at least five times among all models) are labeled.

### 3.4 Evaluation of model fitting

To evaluate the performance of the MSEIR model, we derived two statistical metrics for each of the ten models: *r*^2^ and *root-mean-square error* (*RMSE*). Specifically, we calculated *r*^2^ and *RMSE* for each of the 169 towns by comparing the simulation results with the confirmed cumulative cases in the 50-day period. *r*^2^ assesses whether the MSEIR model captures the trend of the historical data, while *RMSE* quantifies the absolute difference between the model output and the observation.

First, we performed the evaluations across all 169 towns and derived the respective average of the 169 values of *r*^2^ and *RMSE*. Then, the total number of towns with *r*^2^ greater than 0.95 were counted, as shown in Table 2. Overall, all models achieved satisfactory levels of fitting with the historical data in terms of trend monitoring (*r*^2^ > 0.9). The differences in RMSE were more evident. We identified that Model 7 had a better performance than other models in terms of a relatively high *r*^2^ and low *RMSE*.

**Table 2.** Model fitting with the historical data for all towns (*n* = 169).

| Model | *Average $r^2$* | *Average RMSE* | *N ($r^2 > 0.95$)* |
| --- | --- | --- | --- |
| SEIR | 0.919 | 67.869 | 104 |
| 1 | 0.907 | 70.328 | 87 |
| 2 | 0.909 | 72.113 | 92 |
| 3 | 0.910 | 64.059 | 90 |
| 4 | 0.910 | 52.171 | 91 |
| 5 | 0.903 | 61.469 | 81 |
| 6 | 0.907 | 46.957 | 86 |
| 7 | 0.914 | 49.671 | 96 |
| 8 | 0.913 | 60.710 | 94 |
| 9 | 0.910 | 59.457 | 92 |

To further excavate the applicability of the MSEIR model, we further divided the towns into three categories based on the United States Census Bureau (USCB)’s urban-rural classification (USCB 2010): urbanized areas (UAs), urban clusters (UCs), and rural areas (RAs). Then, we evaluated the model fitting for towns under each category, as shown in Table 3. We identified that while UAs and UCs achieved relatively satisfactory levels of model fitting (*r*^2^ > 0.9), RAs or towns with a small population could not be fitted well (*r*^2^ < 0.7). Thus, the MSEIR model is best suited for towns exceeding a certain size (i.e., urban clusters) and must be adjusted for applications for small towns.

**Table 3.** Model fitting with the historical data for towns by category*.

| Model | UAs ($n = 18$) | | | UCs ($n = 135$) | | | RAs ($n = 16$) | | |
| --- | --- | --- | --- | --- | --- | --- | --- | --- | --- |
|  | *Average $r^2$* | *Average RMSE* | *N ($r^2 > 0.95$)* | *Average $r^2$* | *Average RMSE* | *N ($r^2 > 0.95$)* | *Average $r^2$* | *Average RMSE* | *N ($r^2 > 0.95$)* |
| SEIR | 0.985 | 302.367 | 18 | 0.941 | 44.336 | 85 | 0.662 | 2.618 | 1 |
| 1 | 0.972 | 307.222 | 14 | 0.925 | 46.854 | 72 | 0.678 | 1.885 | 1 |
| 2 | 0.978 | 338.683 | 18 | 0.927 | 44.582 | 73 | 0.675 | 4.518 | 1 |
| 3 | 0.979 | 303.672 | 16 | 0.928 | 39.367 | 73 | 0.683 | 2.830 | 1 |
| 4 | 0.977 | 230.632 | 16 | 0.929 | 34.354 | 74 | 0.676 | 1.725 | 1 |
| 5 | 0.973 | 272.269 | 15 | 0.919 | 40.350 | 65 | 0.684 | 2.509 | 1 |
| 6 | 0.976 | 170.620 | 17 | 0.925 | 35.624 | 67 | 0.677 | 3.466 | 2 |
| 7 | 0.981 | 211.781 | 18 | 0.933 | 33.716 | 77 | 0.679 | 1.918 | 1 |
| 8 | 0.983 | 189.952 | 17 | 0.933 | 49.894 | 76 | 0.670 | 6.573 | 1 |
| 9 | 0.980 | 200.609 | 18 | 0.929 | 47.030 | 73 | 0.679 | 5.517 | 1 |

\*The categories are given by USCB (2010) based on population: urbanized areas (UAs) of 50,000 or more people, urban clusters (UCs) between 2,500 and 50,000 people, and rural areas (RAs) of less than 2,500 people. It should be noted that USCB uses a different areal unit for urban-rural classification and is not based on the county subdivision (i.e., town). Thus, the category in this analysis does not suggest the actual urban-rural status of a town.

## 4. Discussion

The proposed MSEIR model applied to meso-scale COVID-19 simulation is among the first to evaluate the development of the pandemic using an administrative unit smaller than a county. By downscaling the analysis to the town level and realizing the model under different social distancing scenarios, the study sheds important insights into COVID-19 studies.

First, meso-scale analysis is of critical importance for revealing the epidemic development after the initial wave of outbreak. When social distancing orders were placed for curbing the early infection, needs for domestic flights or interstate travel were largely suppressed (Gao et al. 2020). New cases of infection would be primarily caused by local spread through short-distance travel. Thus, efforts and policies to contain the COVID-19 development would be most effective by curbing inter- or intra-town travel activities. Incorporating the interaction across townships and deriving their epi curves can help the municipality to leverage resources for preparing for rising contingencies, such as the resurgence of an outbreak in the future. The classical SEIR model and its many extensions, however, lack the capability of simulating the epidemic spread at the meso-scale. This increased spatial granularity to model COVID-19 is the major contribution of the work.

Second, the proposed social distancing framework including the compliance and containment provides quantifiable metrics for COVID-19 studies that attempt to evaluate the effects of social distancing. Since the pandemic is growing at an alarming rate worldwide, existing studies have largely emphasized on the timing (Chinazzi et al. 2020), economic impacts (Atkeson 2020), and the ethical issues (Lewnard and Lo 2020) of social distancing, while the effects on human mobility at the community scale are not well scrutinized. This gap has likely resulted from the lack of detailed mobility data (especially the origin-destination trip data), coupled with the sensitivity of data collection due to the nature of the outbreak and the federal requirement (e.g., the Health Insurance Portability and Accountability Act) for privacy protection. The simulation of the travel activities using the Huff model could help to estimate the regional human movement pattern; and further model calibration will increase the rigor of the estimation (Huff and McCallum 2008).

Lastly, we have found that there are discrepancies in the modeling results across different towns. While the SEIR model has underestimated the majority of the epi curves, it tends to align with towns with a sparse population or located in rural areas, such as Mansfield (Figure 4c) and Wilton (Figure 4f). This alignment is very likely due to the relatively low level of spatial interaction in terms of inter-town travel in these towns. The proposed MERI model works best for towns exceeding a certain size (Table 3) but does not well perform for towns with a complex mobility pattern, such as the city of Hartford (Figure 4b). This result justifies that although towns in a state are subject to the same timing of social distancing orders, the actual policy effects on the residents’ mobility pattern, and consequently, on curbing the epi curves of the pandemic are largely different. Due to this spatial heterogeneity in mobility pattern, which is internally driven by socioeconomic inequities (Bonaccorsi et al. 2020), it is impossible to establish a one-size-fits-all model for COVID-19 analysis for every town in a state. Therefore, we have two recommendations for improving and better understanding the MSEIR model: first, a field survey to solicit people’s daily travel activities (e.g., travel frequency, distance, mode, time, origin-destination) is a necessity to derive the actual mobility pattern between towns in lieu of the model estimation. To fulfill this goal, we have initiated a separate project to investigate people’s actual travel activities during the period of the state easing the lockdown; second, we suggest that local stakeholders employing the modeling results should adopt and prepare for the worst-case scenario (e.g., Model 9) and target towns that may experience the most rapid epidemic growth under all scenarios (e.g., labeled towns in Figure 5). This elevated caution in preparedness can guide the leverage of public health resources towards the most severe situation and the most vulnerable areas.

## 5. Conclusion

The COVID-19 pandemic has posed an unprecedented challenge to the global economy and the health-care system. While modeling COVID-19 by simulating the epic curve has become a growing practice across all disciplines, existing models have not been able to examine the issue at the meso-scale, using a small unit of analysis such as town or census tract, nor have quantified how social distancing may curb the transmission at this scale. The proposed MSEIR model introduces the effects of spatial interaction between towns on the epidemic development. The scenario-based analysis could help policy stakeholders to understand how the compliance with and the containment by social distancing rules regulate people’s travel activities and can help predict how different degrees of policy enforcement would shape the epic curve over different phases of policy implementation. These modeling results have the potential to assist stakeholders with strategical decisions about the timing and expected outcomes of relieving social distancing rules. It should also be noted that with the rapidly evolving epidemic situation and many emerging local policies as countermeasures, the actual epi curves would be very different from what the model predicts. Albeit the tiers of uncertainties, we believe the developed MSEIR model will establish the foundation for community-level assessment and better preparedness for COVID-19.

## Data Availability

Data is available upon request

